# Efficacy and safety of tocilizumab in the management of COVID-19: A systematic review and meta-analysis of observational studies

**DOI:** 10.1101/2021.01.27.21250599

**Authors:** Gollapalle L Viswanatha, CH K V L S N Anjana Male, Hanumanthappa Shylaja

## Abstract

**Background:** This systematic review and meta-analysis was aimed to evaluate the efficacy and safety of tocilizumab (TCZ) in treating severe coronavirus disease 2019 (COVID-19).

**Methods:** The electronic search was performed using PubMed, Scopus, CENTRAL, and Google scholar to identify the retrospective observational reports. The studies published from 01 January 2020 to 30th September 2020. Participants were hospitalized COVID-19 patients. Interventions included tocilizumab versus placebo/standard of care. The comparison will be between TCZ versus standard of care (SOC)/placebo. Inconsistency between the studies was evaluated with *I^2^* and quality of the evidences were evaluated by Newcastle-Ottawa scale.

**Results:** Based on the inclusion criteria there were 24 retrospective studies involving 5686 subjects were included. The outcomes of the meta-analysis have revealed that the TCZ has reduced the mortality (M-H,RE-OR −0.11(−0.18 to −0.04) 95% CI, p =0.001, *I^2^* =88%) and increased the incidences of super-infections (M-H, RE-OR 1.49(1.13 to 1.96) 95% CI, p=0.004, *I^2^*=47%). However, there is no significant difference in ICU admissions rate (M-H, RE-OR −0.06(−0.23 to 0.12), *I^2^*=93%), need of MV (M-H, RE-OR of 0.00(−0.06 to 0.07), *I* = 74%), LOS (IV −2.86(−0.91 to 3.38), *I^2^*=100%), LOS-ICU (IV: −3.93(−12.35 to 4.48), *I^2^*=100%), and incidences of pulmonary thrombosis (M-H, RE-OR 1.01 (0.45 to 2.26), *I^2^*=0%) compared to SOC/control.

**Conclusion:** Based on cumulative low to moderate certainty evidence shows that TCZ could reduce the risk of mortality in hospitalized patients. However, there is no statistically significant difference observed between the TCZ and SOC/control groups in other parameters.

## 1. Introduction

Coronavirus diseases (COVID-19) is a viral disease caused by severe acute respiratory syndrome corona virus-2(SARS-CoV-2) that originated from Wuhan city of Hubei province in China in December 2019[1]. Globally it has caused a significant burden on public health through a drastic increase in the morbidity and mortality rate[2]. The available evidence suggests that most of the infected patients will remain asymptomatic or develop mild symptoms, however nearly 20% of the infected individuals would develop severe pneumonia and respiratory distress syndrome (ARDS) that further progress to cytokine storm syndrome and induced end-organ failure[3]. Interestingly, the United States Food and Drug Administration (USFDA) has approved the drugs such as Remdesivir [4], Bamlanivimab [5], and dexamethasone[6] for the treatment of hospitalized patients with COVID-19. Further, the drugs such as decitabine [7], duvelisib [8], and infliximab [9] are currently under clinical development phase for the treatment of COVID-19. In this context, it is well-known that interleukin-6 (IL-6) is a pleiotropic cytokine that plays a pivotal role in immune-regulation, inflammation, and infection [10, 11]. Noteworthy, the elevated levels of IL-6 in the blood is highly correlated with the mortality rate in the COVID-19 infected patients [12]. The activation of the IL-6 amplifier would induce cytokine storm, a hallmark of dysregulated inflammation, and thus inhibition or blockade of IL-6 amplifier would alleviate cytokine storm in COVID-19 [12].In these lines many studies have reported that TCZ administration could stabilize the health status of COVID-19 patients by improving respiratory functions, reducing CRP levels, and improved health deteriorations due to COVID-19 [12]. Besides, there are multiple case study series, retrospective and prospective study reports available on the therapeutic benefits of TCZ in COVID-19. As of now there are five randomized controlled trials (RCTs) reported on the use of TCZ in COVID-19 (RCT-TCZ-COVID-19 NCT04346355, CORIMUNO-19 NCT04331808, BACC Bay Tocilizumab Trial NCT04356937, COVACTA NCT04320615, REMAP-CAP NCT02735707, and EMPACTA NCT04372186), the low number of subject enrollments In those studies are considered as major limitations and authors have highlighted the need of multicentric RCTs involving a higher number of subjects to determine the safety and efficacy of tocilizumab in COVID-19. In this context, there are several randomized controlled trials registered and under progress to evaluate the clinical benefits of TCZ in alleviating COVID-19 and associated health problems (phase II; NCT04317092, NCT04445272, NCT04377659, NCT04330638, NCT04345445) [13].

With this background, the present study was undertaken to evaluate the clinical benefits of TCZ when administered alone and in combination with standard of care (SOC) and/or placebo in reducing the COVID-19-induced mortality, ICU admissions, MV, LOS, LOS-ICU, super-infections, and pulmonary thrombosis.

## 2. Methodology

A detailed literature search was performed using electronic databases such as PubMed, Science direct, CENTRAL (Cochrane Central Register of Controlled Trials (RCTs), and google scholar to identify the clinical reports (retrospective). The keywords such as ‘Coronavirus disease 2019’ OR ‘Coronavirus infection’ OR ‘Coronavirus’ OR ‘SARS COV-2’ OR ‘nCOV 2019’ ‘Severe acute respiratory syndrome COV 2’ AND ‘Tocilizumab’ OR ‘Interleukin-6 inhibitors’ OR ‘Cytokine storm’ and ‘COVID-19 treatment’ were used. Grey (unpublished) literature was searched in the following trial registries: US National Institutes of Health (NIH; https://clinicaltrials.gov/) and the WHO International Clinical Trials Registry Platform (ICTRP; https://apps.who.int/trialsearch/). Further, the pre-print servers such as Research Square, bioRxiv.org, and medRxiv were also considered while searching the grey literature. Besides, authors have approached the domain experts, seeking their suggestions and inputs in identifying the additional studies (if any) relevant to the topic. The search was not restricted to any publication language or status of the trial. Furthermore, the reference lists of all relevant articles were hand-searched to find additional studies. An example of a search strategy using PUBMED and Google Scholar has been highlighted in **Annexure 1**.

### 2.1. Inclusion criteria

The studies published from 01 January 2020 to 30^th^ September 2020 involving comparison of TCZ group with SOC/control treatment group were included. The studies included in this work involves RT-PCR confirmed cases of COVID-19 (Population), having tocilizumab and corresponding SOC/control as interventions (Intervention), comparison between tocilizumab versus SOC/control (Comparison) for the parameter of interest, the evaluations such as Mortality, ICU admissions, MV, LOS, LOS-ICU, super-infections and pulmonary thrombosis (Outcomes) were included in the study.

### 2.2. Exclusion criteria

1. Studies reporting incomplete data.
2. Single-arm studies.
3. Duplicates, case reports, case series were excluded.
4. *In-vitro* and pre-clinical studies
5. Studies reporting qualitative outcomes without numerical data

The authors of the shortlisted articles were approached through e-mail wherever additional clarification was required. Such as allocation of subjects to control and treatment group, Baseline evaluations, confounding variables, classification of interventions, if there are nay deviations in the intended interventions, measurement of outcomes, data handling, if there is any missing data, reason for missing of data (like selective reporting), treatment details like details of standard of care (SOC) and tocilizumab, the adjustments in the analysis, and so on. The exclusion was executed upon mutual discussion and agreement of all the authors as per exclusion criteria mentioned in the manuscript.

### 2.3 Quality assessment and Risk of Bias analysis for included studies

All the included studies were subjected to the quality assessment using the Newcastle-Ottawa scale and also evaluated for risk of bias using the Cochrane Collaboration tool to assess The Risk of Bias in Non-randomized Studies of Interventions (ROBINS-I).

### 2.4. Parameters

The parameters related to COVID-19 such as Mortality, ICU (intensive care unit) ward admission rate, the need of mechanical ventilation, length of hospital stay (LOS), length of hospital stay in the ICU (LOS-ICU), and the incidences events such as super-infections, fungemia, bacteremia, pneumonia, and pulmonary thrombosis were evaluated as the primary outcomes. The comparison will be between TCZ and standard care/placebo/control.

### 2.5. Article selection, Data extraction, and Analysis

The article selection and data extraction was performed by two reviewers separately based on the inclusion and exclusion criteria listed above. The analysis was carried out at three levels namely on title, abstract and full-text level. Any disagreement was resolved by discussing it with the third reviewer. Two authors have individually extracted the data such as details of participants, methods, interventions, frequency/duration of treatment, outcome measurements, and adverse effects from the included studies. For studies that reported results only in graphical form, numerical values from the graphs were extracted using Adobe® Reader® XI inbuilt measuring tool, version 11.0.06, (Adobe Systems Incorporated, San Jose [California]). Any disagreement was resolved by discussing it with the third reviewer.

### 2.6. Statistical analysis

Review Manager (RevMan, version 5.3; Nordic Cochrane Centre [Cochrane Collaboration], Copenhagen, Denmark; 2014) shall be used to analyze the data. For continuous, variables, inverse variance (IV) was estimated using the random-effects model with a mean difference (MD) or standardized mean difference (SMD) as an effect measure and for the dichotomous variables, the Mantel-Haenszel (M-H) statistic was estimated using a random-effects model with an odds ratio (OR) as the effect measure. Heterogeneity shall be calculated with the *I^2^* statistic. This test estimates the percentage of variation between study results that is due to heterogeneity rather than sampling error. *I^2^* of less than 40% is considered unimportant while that of more than 40% is viewed as moderate to considerable heterogeneity.

## 3. Results

A total of 1425 articles were identified based on the online search, of which 24 articles involving 5676 participants were selected for systematic review and meta-analysis (List of excluded studies based on the full-text screening are given in **Annexure 2**). The PRISMA flow chart of the studies selected is given in **Figure 1**. Only Retrospective studies were selected for the analysis; the characteristics of the included studies are summarized in **Table 1**. In all the included studies TCZ was common and the effect of TCZ was compared with the control group; while in few studies, both standard treatment and TCZ groups had background/previously received either antibiotics, antiviral drugs and corticosteroids, and oxygen which are considered as the standard of care (SOC), and in these studies, the comparison was made between TCZ + SOC versus SOC alone, here the SOC alone is considered as placebo. In the studies where multiple doses of TCZ was used, the response for mid-dose was considered for analysis. In this study, the parameters such as mortality, ICU ward admission rate, need of mechanical ventilation (MV), Length of Hospital Stay (LOS), Length of Hospital Stay in ICU (LOS-ICU) and incidences of events such as super-infections, fungemia, bacteremia, pneumonia, and pulmonary thrombosis were compared between TCZ treatment versus control/SOC groups in COVID-19 positive patients.

**Figure 1.**
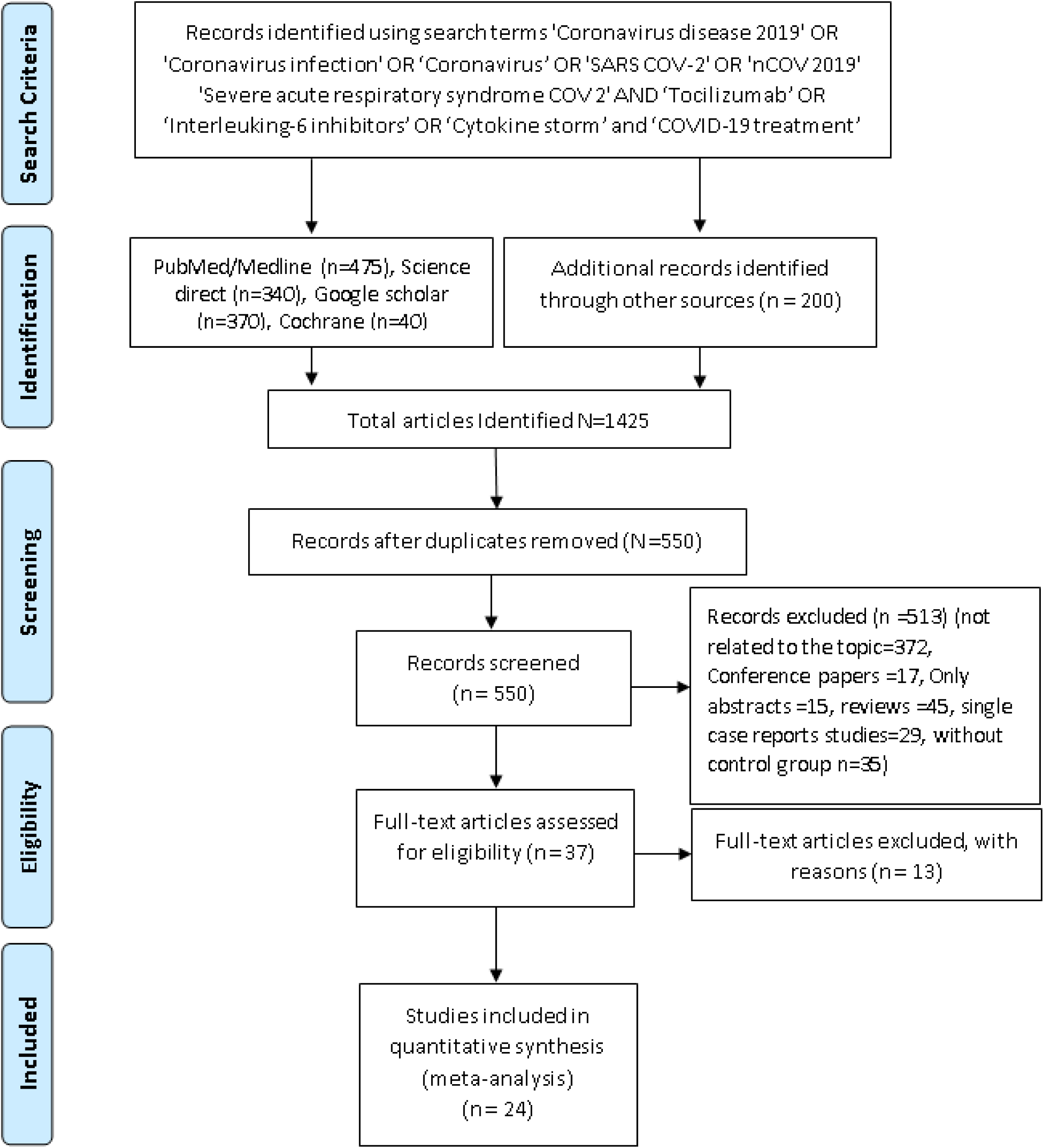
PRISMA Flow chart

### 3.1. Quality assessment and Risk of Bias (RoB)

All the included studies have passed the quality assessment and showed a low risk of bias. The quality assessment and Risk of Bias (RoB) assessment for all the 24 included observational studies are given in **Annexure 3** and **4** respectively.

### 3.2. Efficacy

The improvement in the parameters such as mortality, ICU ward admission rate, need of mechanical ventilation (MV), Length of Hospital Stay (LOS), and Length of Hospital Stay in ICU (LOS-ICU) were considered for evaluating and concluding the efficacy of TCZ compared to control group in COVID-19 positive patients.

#### 3.2.1. Mortality rate

The COVID-19 positive patients treated with TCZ have shown a mortality rate of 24.3 % (448/1841), whereas the control group received SOC has a mortality rate of 31.2% (1079/3454). The outcomes of the meta-analysis has revealed that the TCZ treatment has reduced the mortality rate (Mantel-Haenszel (M-H), random effects Odds ratio (RE-OR) of 0.56 (0.38 to 0.84), at 95% CI, p = 0.005, *I^2^* = 83%) compared to control/SOC. The effect of TCZ on COVID-19 induced mortality of depicted in **Figure 2**.

**Figure 2.**
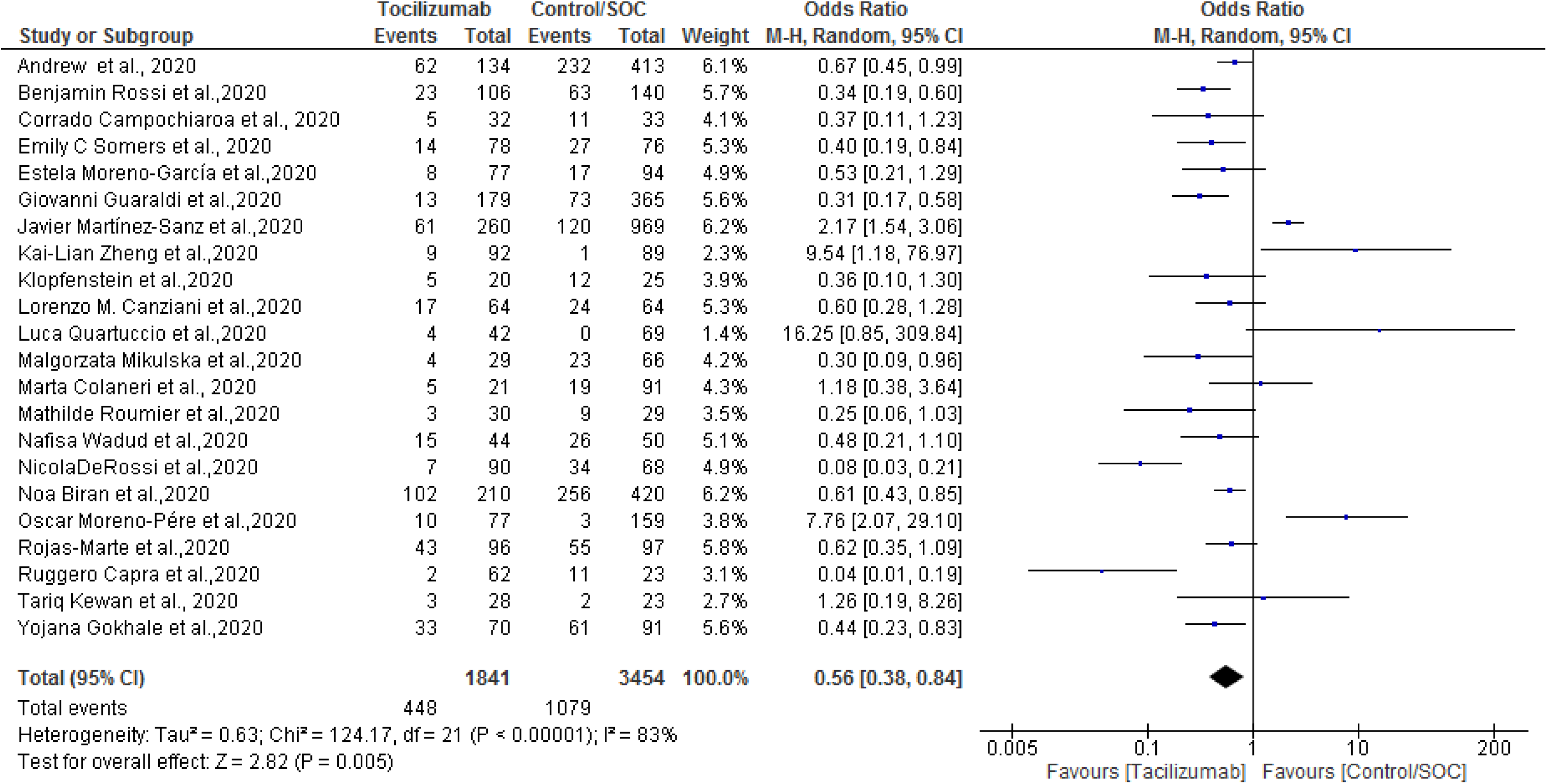
Effect of Tocilizumab on COVID-19 mortality

#### 3.2.2. ICU ward admission rate

Six retrospectives studies involving 542 patients in the TCZ treatment and 1595 patients in the control/SOC, with a total of 2137 COVID-19 positive patients were considered for the analysis. The outcomes of the meta-analysis revealed that there is a statistically significant difference observed between the TCZ and control/SOC treatments in reducing the incidences of ICU ward admission rate (M-H, RE-OR of 0.91 (0.24 to 3.44) at 95% CI, p=0.89, *I^2^* = 94%). The results are given in **Figure 3**.

**Figure 3.**
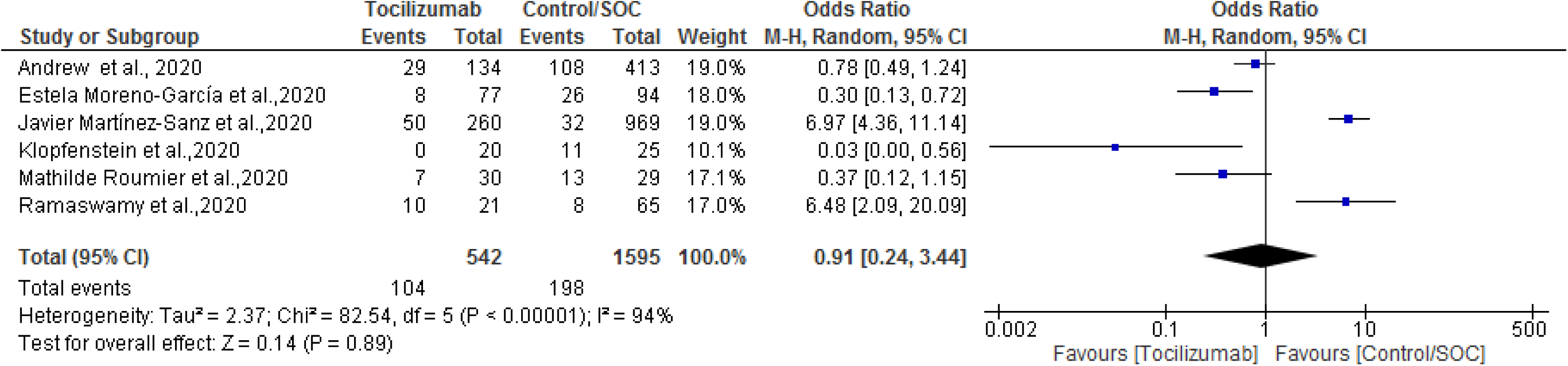
Effect of Tocilizumab on COVID-19 related ICU ward admission rate

#### 3.2.3. need of Mechanical ventilation (MV)

Twelve retrospectives studies involving 756 patients in the TCZ treatment and 1052 patients in the control/SOC, with a total of 1808 COVID-19 positive patients were considered for the analysis. The outcomes have revealed that there is no difference between the TCZ and control/SOC group in the terms of need of mechanical ventilation during the hospitalization (M-H, RE-OR of 1.11 (0.68 to 1.81) at 95% CI, p=0.69, *I^2^* = 63%). The forest plot analysis is depicted in **Figure 4**.

**Figure 4.**
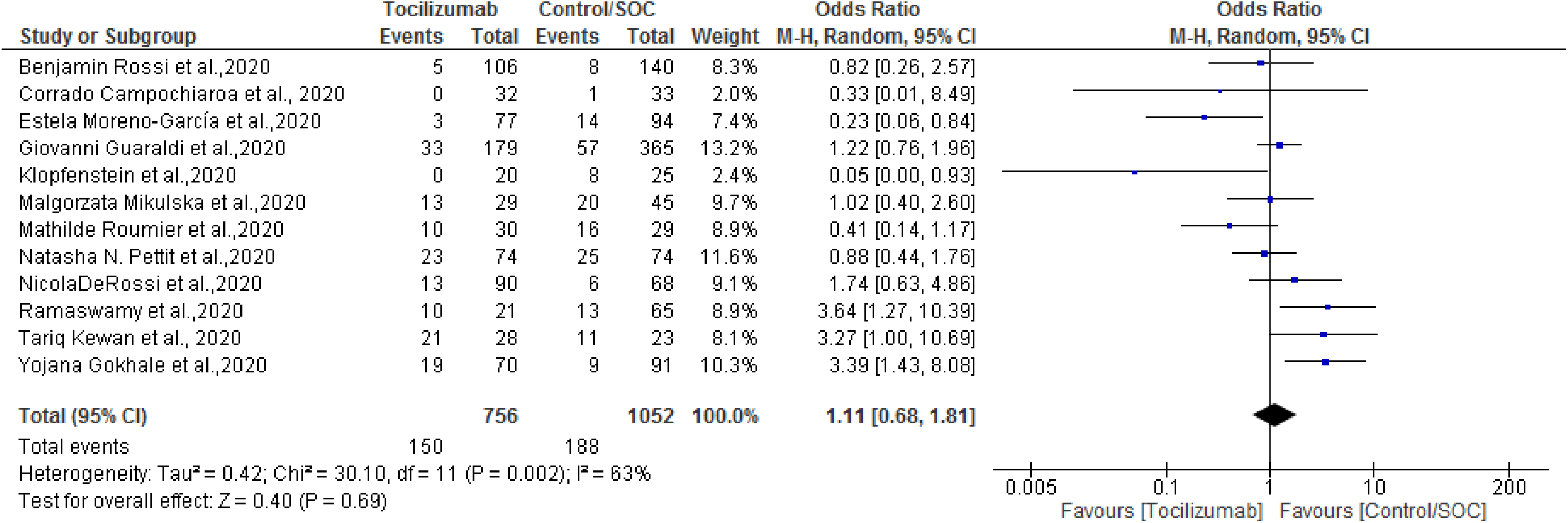
Effect of Tocilizumab on the need of Mechanical ventilation in COVID-19 patients

#### 3.2.4. Effect of TCZ on Length of Hospital Stay (LOS)

The LOS was evaluated by considering the eight retrospective studies comprising of a total of 2030 COVID-19 positive patients, of which 1395 (68.7%) patients were assigned to the TCZ treatment arm and 635 (31.2%) patients were into the control/SOC group. The results of the meta-analysis showed that there was substantial heterogeneity among the included studies (*I^2^*=100%) and there was no statistically significant difference in LOS (days), observed between the TCZ and control/SOC groups (Inverse variance (IV): −2.86 (−0.91 to 3.38) at 95% CI, p=0.37, *I^2^* = 100%). The forest plot analysis for LOS is depicted in **Figure 5**.

**Figure 5.**
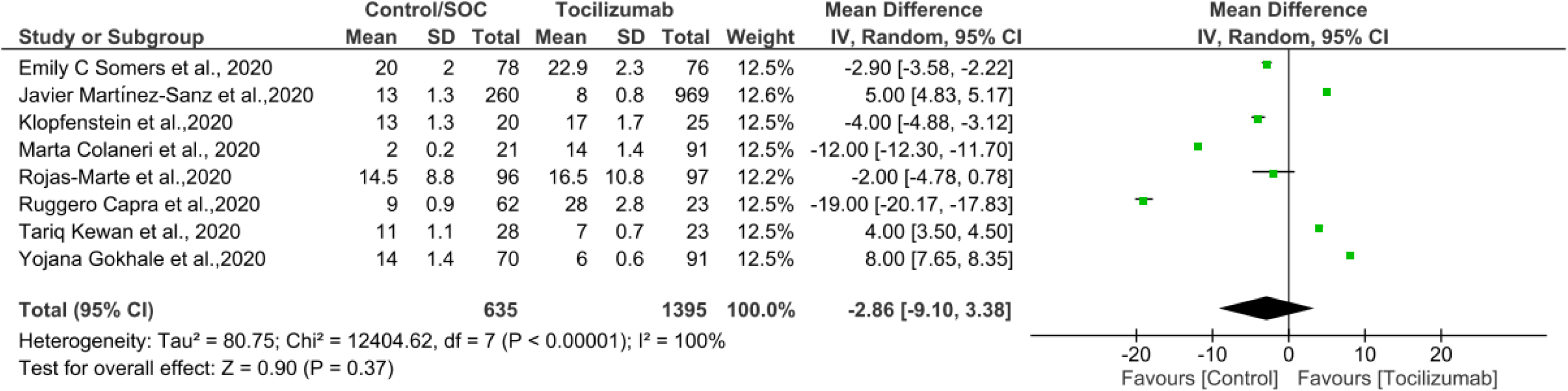
Effect of Tocilizumab on Length of Hospital Stay (LOS) in COVID-19 patients

#### 3.2.5. Effect of TCZ on Length of Hospital stay in ICU (LOS-ICU)

The LOS-ICU was analyzed using three retrospective studies including a total of 1325 COVID-19 positive patients, of which 308 (23.2%) patients were assigned to the TCZ + SOC treatment arm and 1017 (76.8%) patients were into the control/SOC group. The results of the meta-analysis showed that there was substantial heterogeneity among the included studies (I^2^=100%) and there was no statistically significant difference in LOS-ICU (days) observed between the TCZ and control/SOC groups (IV: −3.93 (−12.35 to 4.48) at 95% CI, p=0.36, *I^2^* = 100%). The forest plot analysis for LOS is depicted in **Figure 6**.

**Figure 6.**
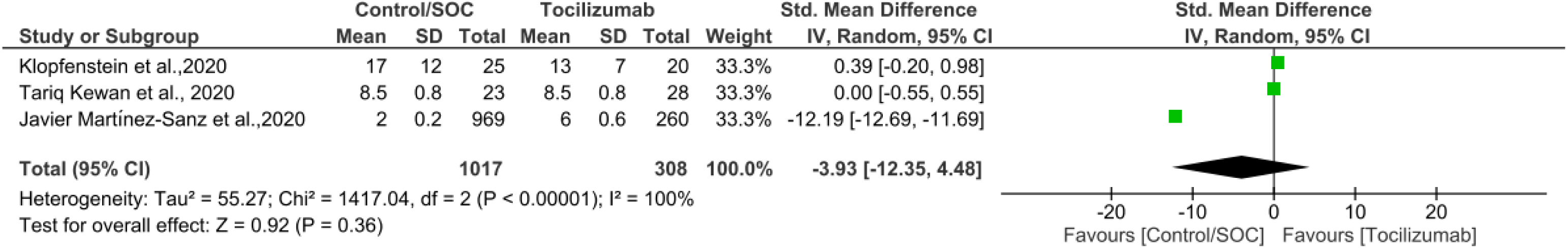
Effect of Tocilizumab on Length of Hospital Stay in ICU (LOS-ICU) in COVID-19 patients

#### 3.2.6. Incidence of Toclimuzab-induced infections

##### 3.2.6 a. Super-infections

The incidence of TCZ-induced super-infections was determined from the eight retrospective studies carried on 1,441 patients out of which 800 (55.5%) were under the control/SOC group and 641(44.4%) patients were considered under the TCZ group.

The meta-analysis revealed that the chances of super-infections are slightly high in the TCZ treated group compared to control/SOC (M-H, RE-OR of 1.81 (1.08, 3.01) at 95% CI, p=0.02, *I^2^* = 60%), however, there is a significant heterogenicity among the studies included for the analysis.

##### 3.2.6 b. Fungemia

The incidence of TCZ-induced fungemia was determined from the three retrospective studies carried on 392 patients, out of which 194 (49.5%) were under the control group and 198(50.5%) patients were assigned under the TCZ group. The meta-analysis revealed that there is no statistically significant difference among the TCZ and control/SOC groups observed in the incidences of fungemia (M-H, RE-OR of 1.73 (0.51, 5.87) at 95% CI, p=0.38, *I^2^* = 0%).

##### 3.1.6 c. Bacteremia

The incidence of TCZ-induced bacteremia was determined from the four retrospective studies carried in 956 patients, out of which 571 (59.3%) patients were under the control/SOC group and 385 (40.2%) patients were assigned under the TCZ group. The outcomes of the meta-analysis showed no difference among the TCZ and control/SOC groups, in the incidences of bacteremia (M-H, RE-OR of 0.92 (0.46, 1.82) at 95% CI, p=0.80, *I^2^* = 35%).

##### 3.2.6 d. Pneumonia

This parameter was analyzed by considering the four retrospective studies carried on 897 patients, out of which 538 (60%) were under the control group and 359(40%) patients were assigned into the TCZ group. The outcomes of the analysis revealed that TCZ treated group has more chances of pneumonia compared to the control/SOC group (M-H, RE-OR of 2.44 (1.50, 3.96) at 95% CI, p=0.0003, *I^2^* = 0%).

. The forest plot analysis of the incidence of TCZ-induced infections are depicted in **Figure 7**.

**Figure 7.**
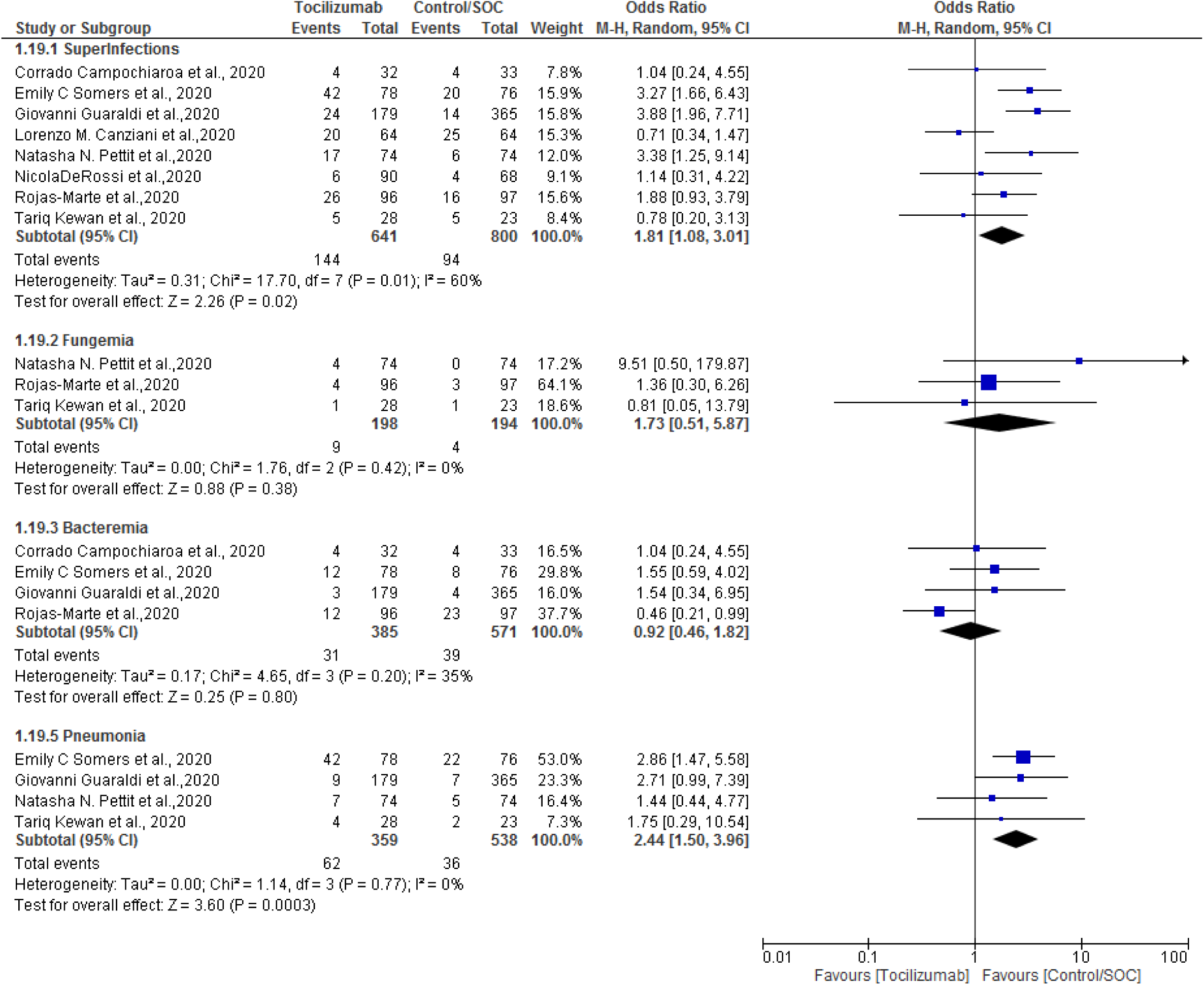
The incidences of Toclimuzab-induced super-infections in COVID-19 patients

#### 3.2.7. Incidence of Toclimuzab-induced pulmonary thrombosis

The incidence of pulmonary thrombosis was determined from the two retrospective studies carried on 193 patients, of which 97 (50.3%) patients were under the control/SOC group and 96(49.7%) patients were assigned under the TCZ group (received at least one dose of TCZ). The outcomes of the analysis revealed that there was no significant difference in the incidences of pulmonary thrombosis between TCZ and control/SOC treated groups (M-H, fixed effect odds ratio (RE-OR) of 1.01 (0.45 to 2.27) at 95% CI, p=0.98, *I^2^* = 0%). The forest plot analysis of the incidence of TCZ-induced pulmonary thrombosis is depicted in **Figure 8**.

**Figure 8.**
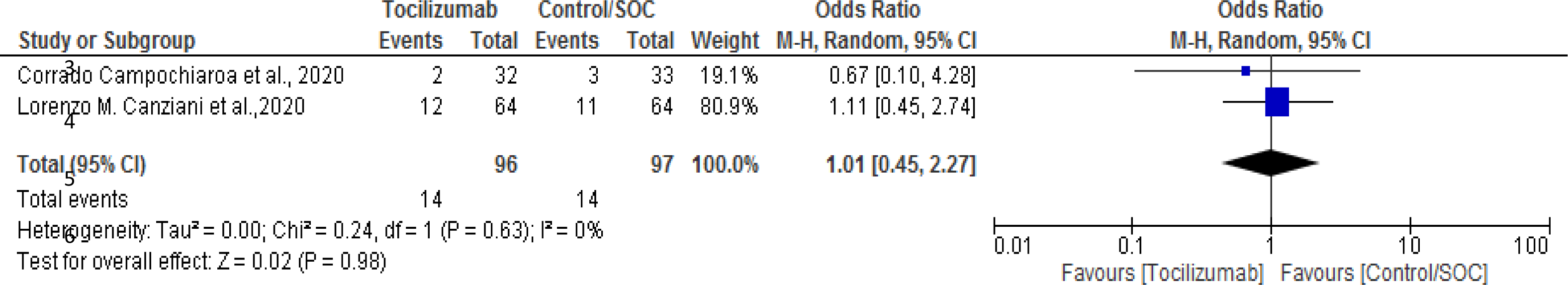
The incidents of Toclimuzab-induced pulmonary thrombosis in COVID-19 patients

## 4. Discussion

This systematic review and meta-analysis were performed to collect, analyze, interpret and conclude the efficacy and safety of TCZ in the treatment of COVID-19 positive subjects. Since the emergence of the pandemic, globally the scientists are in search of a medicine or treatment strategy to combat or manage the pandemic and thereby reduce the social-economic burden throughout the world [14]. Many multi-national organizations, research organizations, and academic researchers are extensively working on developing an effective treatment for COVID-19 [15]. As of now, it has been symptomatically managed by using already existing medications such as antivirals (remdesivir, oseltamivir, etc.), anti-pyretic (paracetamol), anti-histaminic (cetirizine,), antibiotics (cephalosporins), corticosteroids (prednisolone, methylprednisolone), monoclonal antibodies (tocilizumab, imatinib) [16]. In this context, the IL-6 antagonists are considered to have better therapeutic benefits in the symptomatic management of COVID-19, in these lines, TCZ is considered as one of the most commonly used medication to suppress the cytokine storm in COVID-19 patients [17]. There are multiple case series, retrospective, and prospective studies available on the therapeutic role of TCZ in COVID-19. Also, there are few meta-analysis reports published related to the therapeutic use of TCZ in COVID-19. However, this study was focused to provide an overall view on the efficacy and safety of TCZ in COVID-19 patients considering the retrospective studies/reports available on the topic till October 2020. Based on the inclusion and exclusion criteria a total of 24 retrospective studies were selected for this study, comprising of 5686 COVID-19 positive patients. The parameters such as mortality, ICU ward admission rate, need of Mechanical ventilation, Length of Hospital Stay, Length of Hospital stay in ICU, Incidence of super-infections such as fungemia, bacteremia, pneumonia, and pulmonary thrombosis were considered for systematic review and meta-analysis.

The available literature favors the benefit of TCZ in minimizing the COVID-19 induced mortality [18, 19, 20, 21]. In this study, 22 included studies have reported mortality as a parameter of which 16 studies have supported the use of TCZ in minimizing the mortality, and 6 studies have shown results in the favor of control. The outcome of the meta-analysis revealed that the administration of TCZ could benefit the COVID-19 positive patients in minimizing the COVID-19 induced mortality.

Further, among the hospitalized COVID-19 positive patients about 5-10% population requires ICU admission [22, 23]. In this regard, some of the available reports have supported the use of TCZ in reducing the incidences of ICU admission [22, 24, 25]. However, the outcomes of the present meta-analysis showed that there is no significant difference between the TCZ and control/SOC groups in the incidences of ICU admission among the COVID-19 infected patients. Moreover, about 89.9% of ICU cases and 20.2% of hospitalized COVID-19 positive patients require mechanical ventilation (Invasive and/or Non-invasive) [26], and the role of TCZ in reducing the need for MV has been evaluated in multiple studies [27, 28, 29,30, 31]. In the present study, 12 retrospective studies reporting the MV as a parameter were included, of these 12 studies, 7 studies have reported in favor of TCZ, 5 studies have reported in the favors of control/SOC and 1 study was neutral. The outcomes of the meta-analysis revealed that the need for MV is the same between TCZ and control/SOC treated groups.

Besides, length of stay (LOS and LOS-ICU) is the one of important parameters considered for evaluating the efficacy in COVID-19 patients. As per the available data, the median LOS was found to be 4 to 21 days (outside China) and median LOS-ICU was found to be 4 to 19 days (outside China) [32]. In this regard, 8 studies reporting LOS and 3 studies reporting LOS-ICU were included in the present study. On the overview, the TCZ treatment has shown benefit in minimizing the LOS and LOZ-ICU compared to control/SOC. However, there was no statistically significant difference observed between the TCZ and control/SOC groups, due to the significant heterogenicity (*I^2^* = 100%) associated with the included studies.

On the other hand, the safety of TCZ is the prime concern while administering to COVDI-19 patients. The available literature suggests that TCZ administration can cause adverse (AEs) and serious adverse events (SAEs) such as super-infections, fungemia, bacteremia, pneumonia, pulmonary thrombosis, and so on [30, 33, 34, 35, 36, 37]. Therefore, the incidences of events such as these adverse events were compared between the TCZ and control/SOC treated groups using the 9 included studies. The outcomes of the meta-analysis revealed that the TCZ administration has higher chances of producing the events such as super-infections, fungemia, bacteremia, pneumonia, and pulmonary thrombosis compared to control/SOC.

Further, we found that there are 7 systematic reviews and meta-analyses published based on observational studies on the role of tocilizumab in the treatment of COVID-19 [45, 46, 47, 48,49, 50, 51]. However, a meta-analysis published by Aziz et al [45] has a close association with this meta-analysis. Aziz et al., included 23 studies involving 6279 patients, and the parameters such as mortality, need for mechanical ventilation, ICU admission, and secondary infections were considered as parameters [45]. However, we have included additional parameters such a LOS, LOS-ICU, role of TCZ treatment on incidences of super-infections, and also evaluated the incidences of TCZ-induced pulmonary thrombosis. In addition, our analysis has a greater number of studies in the parameters like mortality and the need for mechanical ventilation. Lastly, in the conclusion section Aziz et al., have stated that TCZ treatment has the potential to decrease the mortality rate in severe COVID-19 patients without causing a significant increase in the infection rate [45].

Commenting on other meta-analysis works, Zhao et. al, have performed a meta-analysis including 10 studies, comprising 1675 patients. Mortality, admission to ICU, safety, and efficacy were considered for analysis. This study has concluded that TCZ treatment could reduce mortality significantly compared to control/SOC in severe COVID-19 patients [46]. Further, the meta-analysis performed by Liu et. al., has included 28 studies consisting of 991 COVID-19 confirmed patients receiving TCZ. They have concluded that TCZ administration has reduced the death rate in severe COVID-19 cases [47]. Moreover, Surjit Singh et. al have performed a meta-analysis including a total of 13 observational studies comprising 2750 patients. Based on the detailed analysis they have concluded that there is a 46% decrease in mortality rate, and a 66% decrease in the progression of diseases in TCZ treated group compared to the SOC group [48]. Besides, based on the meta-analysis of 16 studies Boregowda et. al. have concluded that the addition of TCZ to the standard regimen could reduce the mortality in severe COVID-19 patients [49]. On the other hand, Kotak S et al have carried a meta-analysis including 13 studies consists of a total of 766 patients. Based on the observations, authors have concluded that TCZ is safe and effective in reducing mortality among critically ill COVID-19 patients. However, this study has very limited numbers of observations. However, a systematic review performed by Lan et al. has concluded that the available pieces of evidence are not strong enough to derive a conclusive decision about the benefit of TCZ in treating COVID-19 and associated health complications. They have included 7 retrospective studies comprising of 592 adult COVID-19 patients. [51].

Basides, there is a meta-analysis work published by Tleyjeh et al., wherein authors have included five RCTs of tocilizumab related to its benefits in COVID-19. The outcomes of the meta-analysis (based on RCTs) concludes that TCZ did not reduce the short-term mortality, and cumulative evidences suggest that there is reuction in risk of mechanical ventilation. While, there was difference in the risk of infections or adverse events between the TCZ and SOC groups [52]. The pooled estimated of meta-analysis of RCTs Overall, based on the meta-analysis of moderately-certain evidence we can conclude that the administration of TCZ would reduce the risk of mortality, and however, there is no much difference observed between the TCZ and SOC/control groups in other parameters such as ICU admission rate, need of mechanical ventilation and length of hospital stay (ICU and Non-ICU). On the other hand, TCZ treated subjects possess higher chances of adverse events like super-infections, fungemia, bacteremia, pneumonia, and pulmonary thrombosis compared to the control/SOC group.

## 5. Conclusions

This meta-analysis was performed using retrospective clinical reports on the use of TCZ in COVID-19, and based on the outcomes of the meta-analysis we can conclude that administration of TCZ would reduce the risk of mortality, and however, there is no much difference observed between the TCZ and SOC/control groups in other parameters such as ICU admission rate, need of mechanical ventilation and length of hospital stay (ICU and Non-ICU). On the other hand, TCZ treated subjects possess higher chances of super-infections and pneumonia compared with SOC/control group. All the included studies have passed the quality assessment and showed a low risk of bias. However, the major limitation of this study is the significant heterogenicity observed in the outcomes due to multiple confounding factors, and hence there is a need for multi-centric randomized trials involving a large COVID-19 patient population, proper adjustment of confounders like SOC medications to determine the potential therapeutic role of TCZ in mitigating COVID-19 and associated health complications.

## 6. Limitations

Our meta-analysis has many limitations. Firstly, all the included articles are observational studies and the observational studies are not as robust as randomized controlled studies. Secodnly, there was a moderate to significant heterogenicity observed in the outcomes of the various parameters evaluated. Interestingly, the observed heterogenicity between the included studies is due to multiple reasons as highlighted below. All the included studies are observational, and the confounders associated with one or two included studies may also influence the outcomes to a large extent. Besides, since there was no specific treatment available for COVID-19 (before the approval of vaccine), the therapies are dynamically evolving with each passing day, therefore the comparator group or standard of care (SOC) or control group has varied significantly from study to study, this is also one of the potential causes for heterogenicity observed. Lastly, there was no standard treatment regimen available for tocilizumab use in the included studies, like there is the difference in dosage regimen (strength and number of doses), route of administration, time of administration, age, and gender difference. These are some of the possible reasons for significant heterogenicity observed in the outcomes of the present study.

## Data Availability

All data related to the manuscript is available in the manuscript itself.

## Annexure 1

### 1. Search strategy using PUBMED

1.Search strategy using PUBMED
| SI No. | Search Terms | Results |
| --- | --- | --- |
| 1 | 'Coronavirus disease 2019' OR 'Coronavirus infection' OR 'Coronavirus' OR 'SARS COV-2' OR 'nCOV 2019' 'Severe acute respiratory syndrome COV 2' | 112565 |
| 2 | 'Tocilizumab' OR 'Interleukin-6 inhibitors' OR 'Cytokine storm' OR 'COVID-19 treatment' | 13178 |
| 3 | 1 AND 2 | 613 |
| 4 | 3 NOT ('Meta-analysis' OR 'Practice guideline' OR 'Systematic review' OR) | 550 |
| 5 | 4 NOT ('Newsletters' OR 'Commentaries' OR 'Opinions' OR 'Editorial' OR 'letter to the editor' OR 'Short survey') | 475 |

### 2. Search strategy using GOOGLE SCHOLAR

| SI No. | Search Terms | Results |
| --- | --- | --- |
| 1 | 'Coronavirus disease 2019' OR 'Coronavirus infection' OR 'Coronavirus' OR 'SARS COV-2' OR 'nCOV 2019' 'Severe acute respiratory syndrome COV 2' | 2013421 |
| 2 | 'Tocilizumab' OR 'Interleukin-6 inhibitors' OR 'Cytokine storm' OR 'COVID-19 treatment' | 18252 |
| 3 | 1 AND 2 | 578 |
| 4 | 3 NOT ('Meta-analysis' OR 'Practice guideline' OR 'Systematic review' OR) | 441 |
| 5 | 4 NOT ('Newsletters' OR 'Commentaries' OR 'Opinions' OR 'Editorial' OR 'letter to the editor' OR 'Short survey') | 370 |

## Annexure 2. List of excluded papers based on full-text evaluation

| Sl. No. | First author-name | Title | Reason for exclusion |
| --- | --- | --- | --- |
| 1 | Betul Borku et al., | Tocilizumab challenge: A series of cytokine storm therapy experiences in hospitalized COVID-19 pneumonia patients | 1.No control group |
| 2 | Federico Alberici et al., | Management of Patients on Dialysis and With Kidney Transplantation During the SARS-CoV-2 (COVID-19) Pandemic in Brescia, Italy | 1.No numerical data reported |
| 3 | Marcus R. Pereira et al., | COVID-19 in solid organ transplant recipients: Initial report from the US epicenter | 1. Patients have a history of solid organ transplant<br>2.No control group |
| 4 | Marfella et al., | Negative impact of hyperglycemia on tocilizumab therapy in Covid-19 patients | 1. Evaluations related to glycaemic control in the diabetic population<br>2. No control group<br>3. No numerical data |
| 5 | Maria Mazzitelli et al., | Use of subcutaneous tocilizumab in patients with COVID-19 pneumonia | 1.No numerical data<br>2.No control group |
| 6 | Nahéma Issa et al., | Feasibility of Tocilizumab in ICU patients with COVID-19 | 1.No control group<br>2. Only biochemical parameters are considered, which are out of the scope of the present review. |
| 7 | Nan Yu et al., | Clinical features and obstetric and neonatal outcomes of pregnant patients with COVID-19 in Wuhan, China: a retrospective, single-center, descriptive study, March 24, 2020: 30176-6.<br>[http://doi.org/10.1016/S1473-3099(20)30176-6](http://doi.org/10.1016/S1473-3099%2820%2930176-6). | 1.Case series without a parallel control |
| 8 | Pan Luo et al., | Tocilizumab treatment in COVID-19: A single-center experience | 1. Case series without a control group.<br>2. Only CRP and IL-6 were considered as parameters, which are not out of the scope of the present review. |
| 9 | Patel K et al., | Use of the IL-6R antagonist tocilizumab in hospitalized COVID-19 patients. | 1. No control group |
| 10 | Şiran Keske et al., | Appropriate use of tocilizumab in COVID-19 infection | 1. No control group |
| 11 | Timothy et al., | Tocilizumab for severe COVID-19 pneumonia: Case series of 5 Australian patients | 1. Case series without parallel control. |
| 12 | Tomasiewicz et al., | Tocilizumab for patients with severe COVID-19: a retrospective, multi-center study | 1. No control group<br>2. The parameters evaluated are out of the scope of the present review. |
| 13 | Xu X et al., | Effective treatment of severe COVID-19 patients with tocilizumab. | 1. No control group<br>2. The parameters evaluated are out of the scope of the present review |

## Annexure 3. Quality assessment of Included papers by Newcastle-Ottawa scale (NOS)

| Sl. No | Included Studies | Selection |  |  |  | Comparability | Outcome |  |  | Quality score | Study rating |
| --- | --- | --- | --- | --- | --- | --- | --- | --- | --- | --- | --- |
|  |  | Representativeness of the exposed cohort | Selection of non-exposed cohort | Ascertainment of exposure | Demonstration that outcome of interest was not present at the start of the study | Comparability of the cohorts on the basis of design or analysis | Assessment of outcome | Was follow up long enough for outcomes to occur | Adequacy of follow up cohorts |  |  |
| 1 | Andrew IP et al., 2020 [18] | 1 | 0 | 1 | 1 | 1 | 1 | 1 | 0 | 6 | Good |
| 2 | Biran N et al.,2020[44] | 1 | 1 | 0 | 1 | 1 | 0 | 1 | 1 | 6 | Fair |
| 3 | Campochiaro C et al., 2020 [33] | 1 | 0 | 1 | 1 | 1 | 1 | 1 | 1 | 7 | Good |
| 4 | Canziani LM et al.,2020[36] | 1 | 0 | 1 | 1 | 1 | 1 | 1 | 1 | 7 | Fair |
| 5 | Capra R et al.,2020[21] | 1 | 1 | 1 | 1 | 1 | 1 | 1 | 1 | 8 | Good |
| 6 | Colaneri M et al., 2020 [39] | 1 | 1 | 1 | 1 | 1 | 1 | 0 | 0 | 6 | Fair |
| 7 | De Rossi N et al.,2020 [41] | 1 | 0 | 1 | 1 | 1 | 0 | 1 | 1 | 6 | Fair |
| 8 | Gokhale Y et al.,2020 [31] | 1 | 1 | 1 | 1 | 1 | 1 | 1 | 0 | 7 | Good |
| 9 | Guaraldi G et al.,2020[28] | 1 | 0 | 1 | 1 | 1 | 1 | 0 | 1 | 6 | Fair |
| 10 | Kewan T et al., 2020 [30] | 1 | 0 | 1 | 1 | 1 | 1 | 0 | 1 | 6 | Fair |
| 11 | Klopfenstein T et al.,2020[25] | 1 | 1 | 1 | 1 | 1 | 1 | 0 | 1 | 7 | Good |
| 12 | Martínez-Sanz J et al.,2020 [23] | 1 | 1 | 1 | 1 | 1 | 1 | 0 | 1 | 7 | Good |
| 13 | Mikulska M et al.,2020 [29] | 1 | 1 | 1 | 1 | 1 | 1 | 1 | 1 | 8 | Good |
| 14 | Moreno-García E et al.,2020 [ 22] | 0 | 1 | 1 | 1 | 1 | 1 | 1 | 1 | 7 | Good |
| 15 | Moreno-Pérez O et al.,2020 [42] | 1 | 1 | 1 | 1 | 1 | 0 | 0 | 1 | 6 | Fair |
| 16 | Pettit NN et al.,2020 [37] | 1 | 0 | 1 | 1 | 0 | 1 | 1 | 1 | 6 | Poor |
| 17 | Quartuccio L et al.,2020[38] | 1 | 1 | 1 | 0 | 1 | 0 | 1 | 1 | 6 | Fair |
| 18 | Ramaswamy M et al.,2020 [43] | 0 | 1 | 1 | 0 | 1 | 1 | 1 | 1 | 6 | Fair |
| 19 | Rojas-Marte G et al.,2020 [34] | 1 | 0 | 1 | 0 | 1 | 1 | 1 | 1 | 5 | Fair |
| 20 | Rossi B et al.,2020[19] | 0 | 1 | 0 | 1 | 1 | 1 | 1 | 1 | 6 | Good |
| 21 | Roumier M et al.,2020 [24] | 1 | 0 | 1 | 1 | 0 | 1 | 1 | 1 | 6 | Poor |
| 22 | Somers EC et al., 2020 [35] | 0 | 1 | 1 | 0 | 1 | 1 | 1 | 0 | 6 | Fair |
| 23 | Wadud N et al.,2020 [40] | 1 | 0 | 1 | 1 | 1 | 0 | 1 | 1 | 6 | Fair |
| 24 | Zheng KL et al.,2020 [20] | 1 | 1 | 1 | 1 | 1 | 0 | 1 | 1 | 7 | Good |

## Annexure 4. Risk of Bias assessment for Included studies using ROBINS-I (Risk of Bias In Non-randomised Studies of Interventions) (Sterne Jonathan et al., 2016)

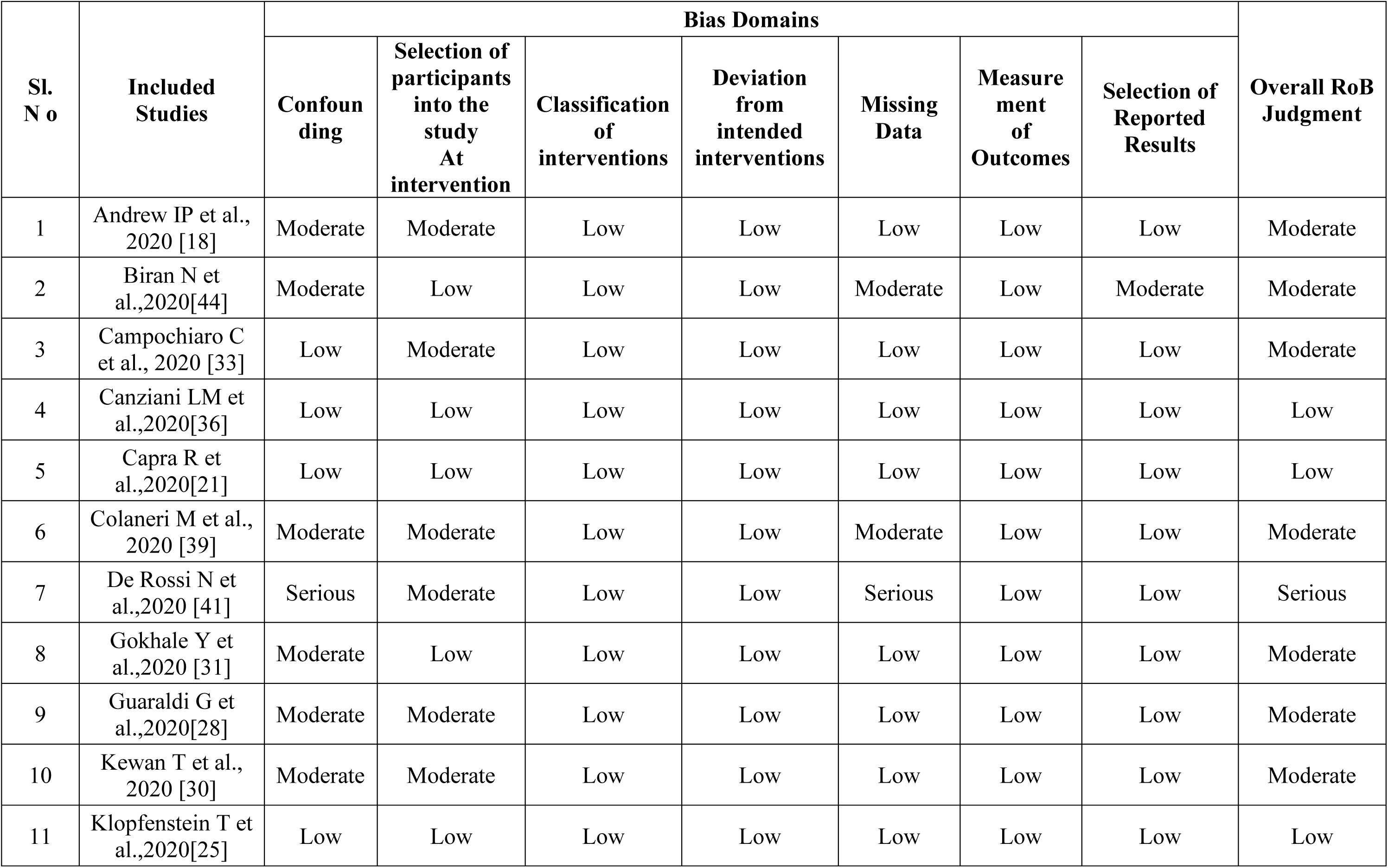
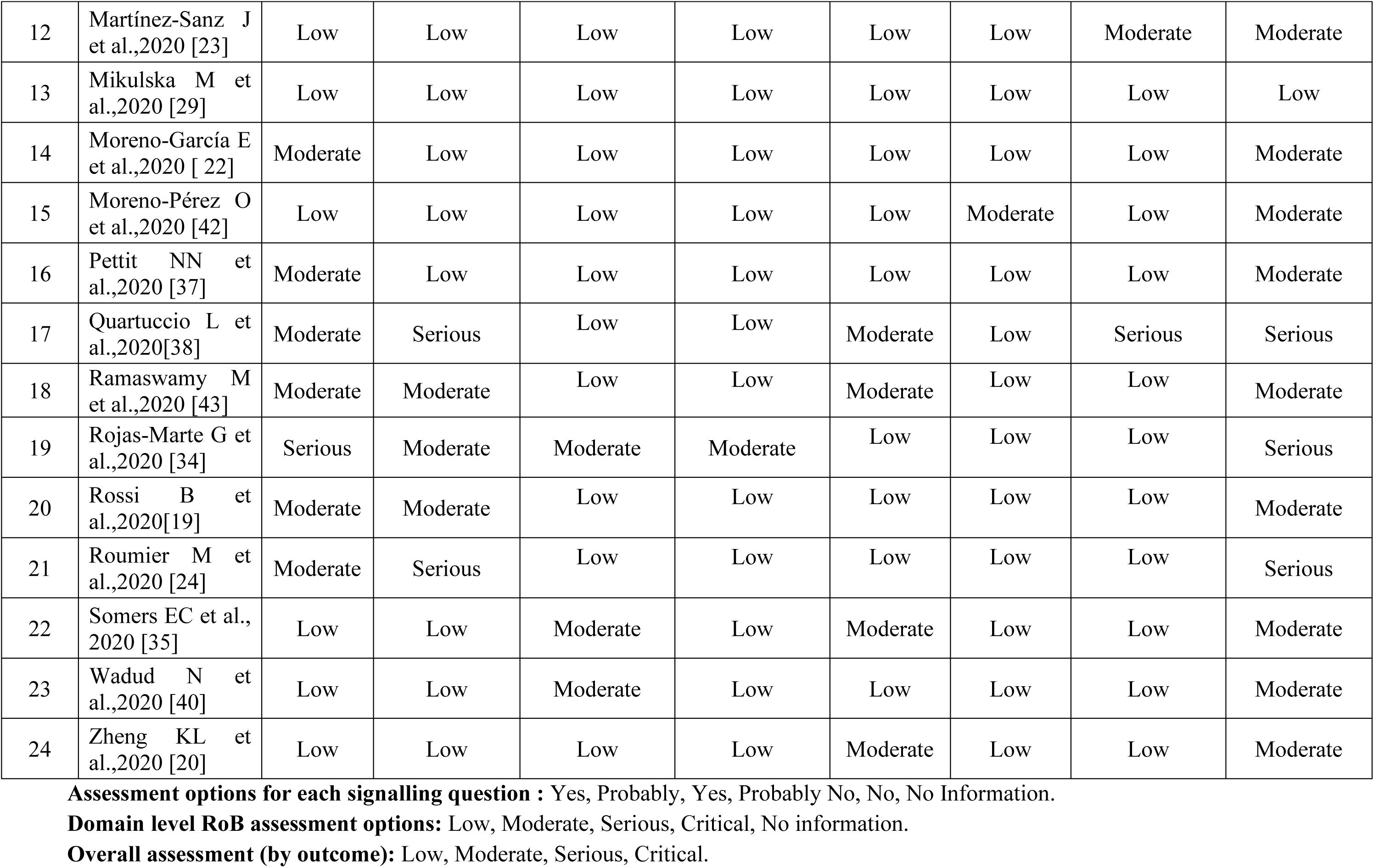
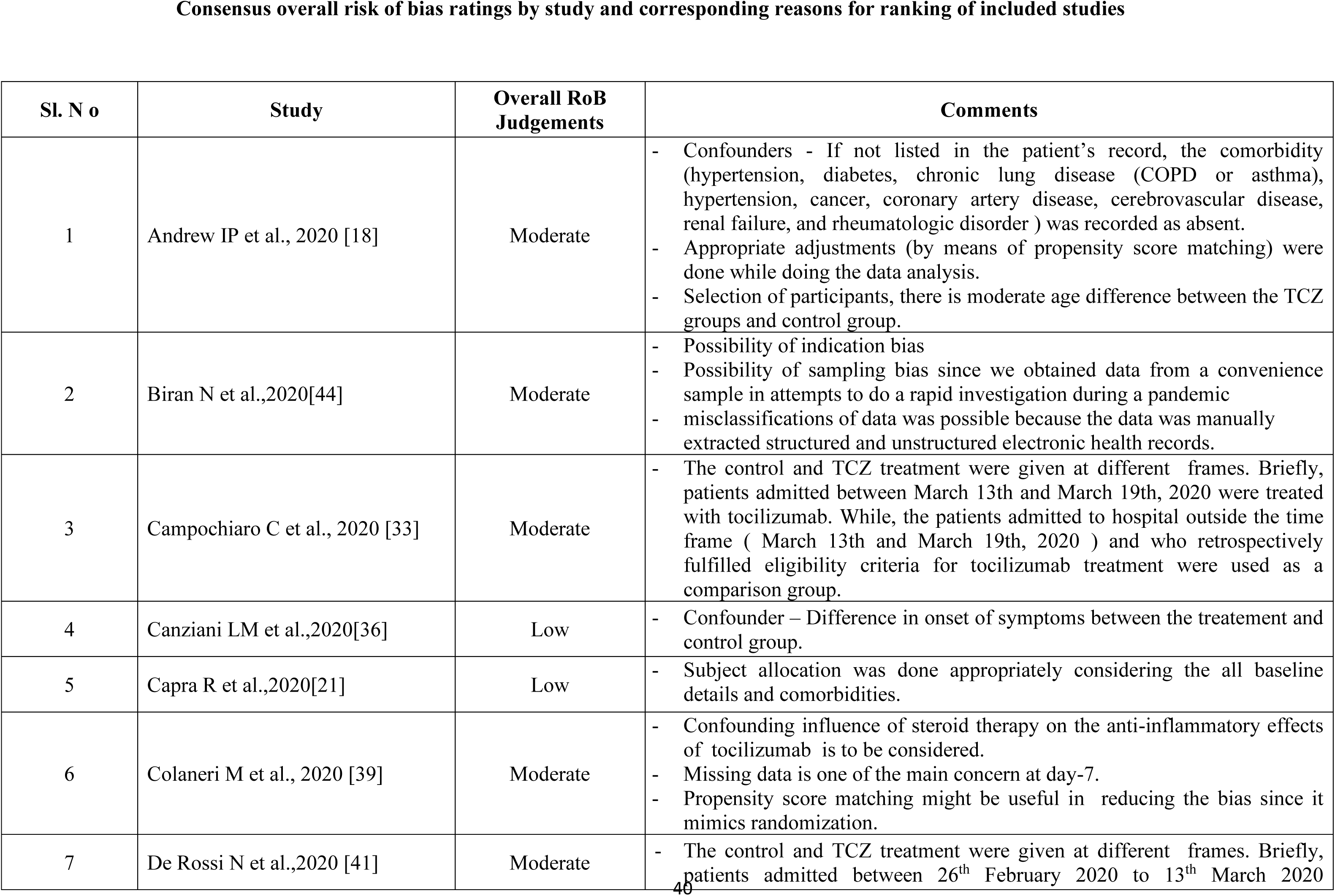
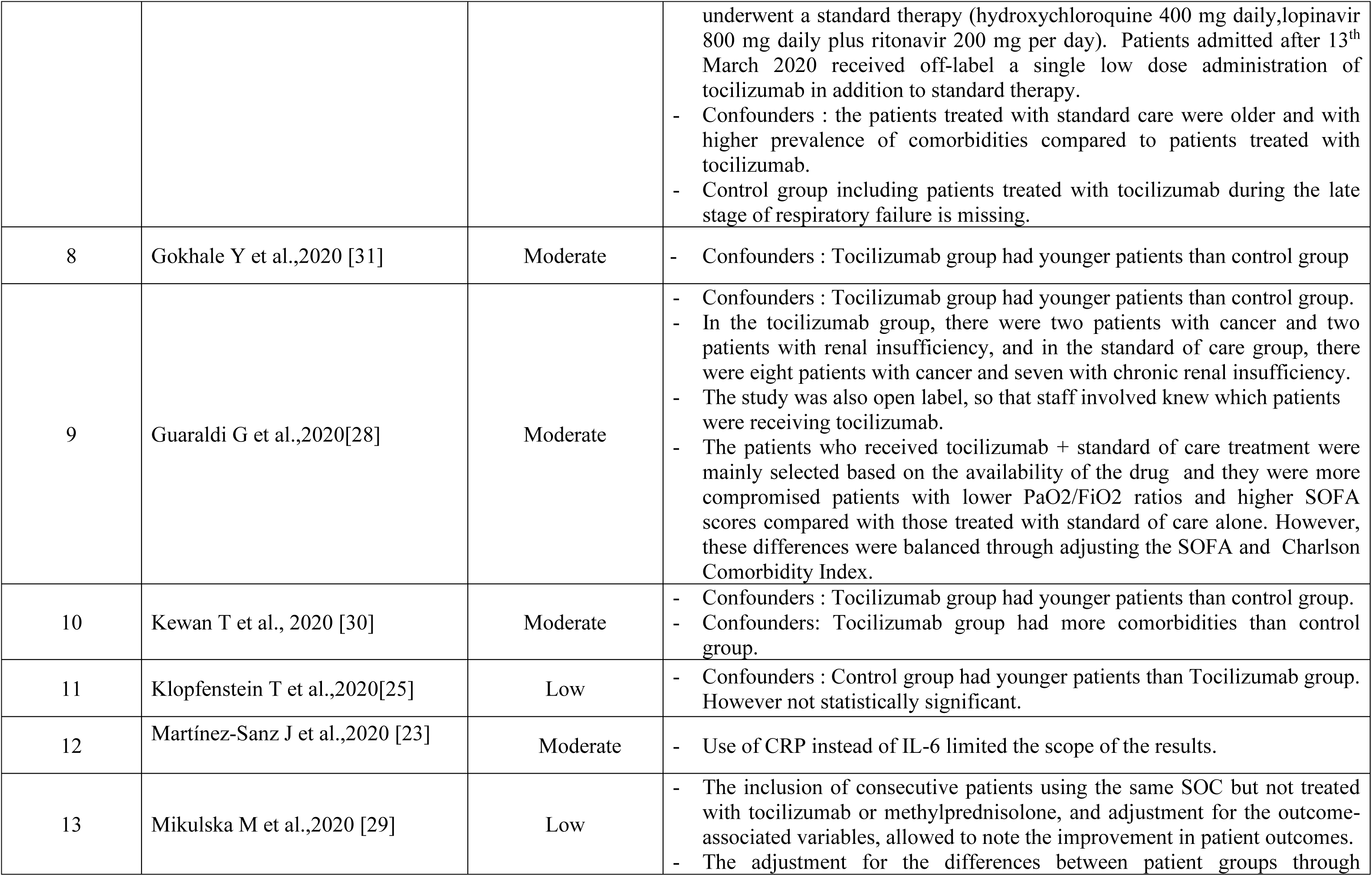
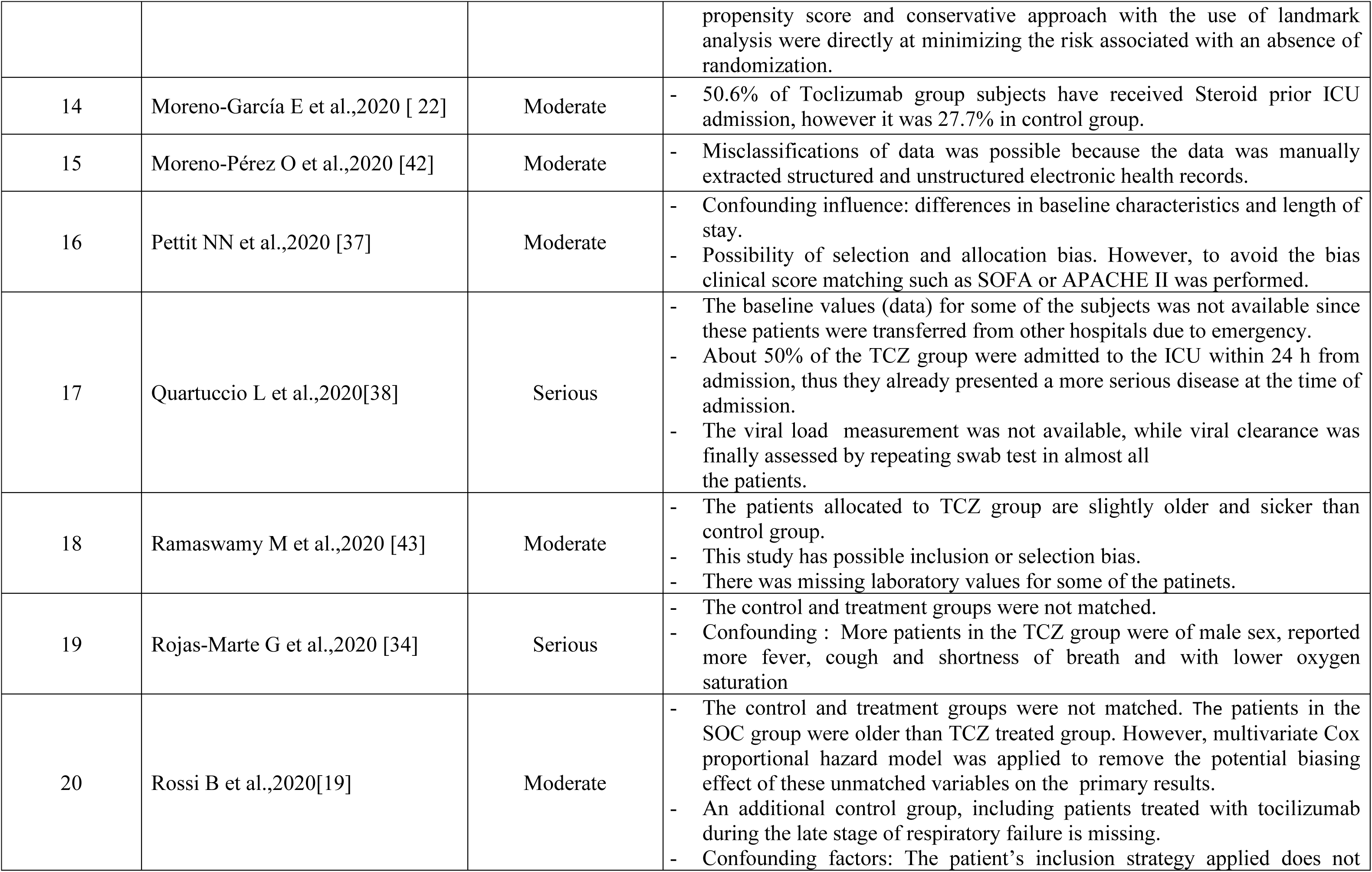
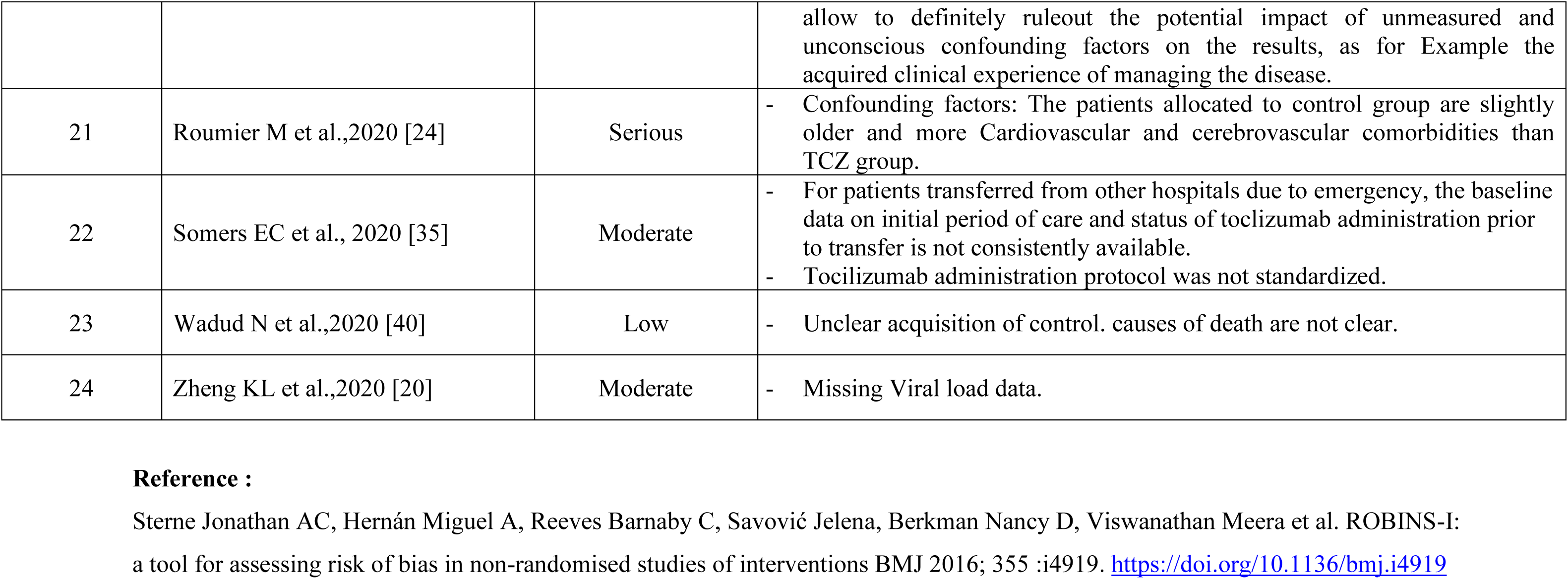

